# Covid19, 2020 - On the number of infected cases and the effective reproduction rate

**DOI:** 10.1101/2020.06.13.20130468

**Authors:** Isaac Meilijson

## Abstract

The SIR differential equations in Epidemiology are re-examined in the context of Covid19, 2020. The number of recovered cases is calibrated in time, and the number of infected cases is derived by a linear regression equation. Emphasis is placed on the adaptive assessment of the effective reproduction rate *R*_0_, especially in countries with under-reported recovered cases. Analysis is applied to the Covid19 2020 data of a number of countries, as reported by the Johns Hopkins University data repository.

## 1 Introduction

The SIR (Susceptibles, Infected, Removed) model introduced by Kermack & McKendrick in 1927 [7] for the progress of an epidemic describes the interdependence between the cumulative number *X*(*t*) of affected cases, the cumulative number *R*(*t*) of removed cases (dead or recovered) and the ensuing current number of infected cases *Y* (*t*) = *X*(*t*) − *R*(*t*). The version of this model to be applied in the current report conforms with the assumption (henceforth the SIR “*γ*-equation”)

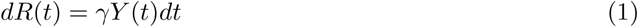

that the daily number of new removed cases Δ*R*(*t*) constitutes a fixed proportion *γ* of the number of currently infected cases *Y* (*t*), and with the assumption that the daily number of new affected cases Δ*X*(*t*) is proportional to the product of *Y* (*t*) by some dimensionless function representing the fraction of susceptible cases in the population, such as 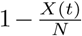, where *N* is some effective population size. However, no such simple mathematical function, with constant *N*, could play this role since the effective reproduction rate 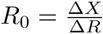 (the driving wheel of epidemic management - the attempt to keep *R*_0_ below 1), is monitored and partially controlled as a non-monotone stochastic process.

In this report, the cumulative number *X*(*t*) of affected cases and *D*(*t*) of dead cases will be taken at face value, as reported. As for the use given to the reported number *REC*(*t*) of recovered cases, the actual cumulative number of recovered cases by day *t* will be considered to be *REC*(*t* + *δ*) where the delay by *δ* days will make the infected cases *Y* (*t*) = *X*(*t*) ™ *D*(*t*) − *REC*(*t* + *δ*) and removed cases *R*(*t*) = *D*(*t*) + *REC*(*t* + *δ*) satisfy the *γ*-equation (1) quite accurately. As will become apparent in most countries used as illustration, a regression analysis to replace this (modified but yet empirical) *Y* by a version *Ŷ* satisfying the *γ*-equation to perfection leaves *Y* practically unchanged. This derived number *Ŷ* of infected cases will play the role of infected cases throughout this study.

The parameter *γ*, estimated as a by-product of this procedure, will be seen to be generally in the narrow range between 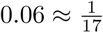 and 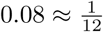. The meaning of *γ* is perhaps better represented by 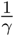, the mean number of days during which an infected individual is contagious.

A number of countries, among these Belgium, France, USA and to a lesser extent Italy, heavily under-report the number of recovered cases. With removed and infected cases unknown, the assessment of the effective reproduction rate *R*_0_, and its crossovers of 1, are out of reach. By postulating some central value of *γ*, a representation of *R* and *Ŷ* satisfying (1) will be obtained, from which *R*_0_ can be monitored.

The data analyzed in this report is taken from the COVID-19 Data Repository of the Center for Systems Science and Engineering (CSSE) at Johns Hopkins University, from January 22, 2020 to October 18, 2020. Data consist of daily triples (*X*_1_, *REC*_1_, *D*_1_), (*X*_2_, *REC*_2_, *D*_2_), …, (*X*_*n*_, *REC*_*n*_, *D*_*n*_) of cumulative numbers of affected, recovered and dead cases respectively. As explained above, the cumulative number of removed cases should in principle be taken as *R*_*t*_ = *REC*_*t*_ + *D*_*t*_ and the number of currently infected cases as *Y*_*t*_ = *X*_*t*_ *− R*_*t*_. However, these statistics fail generally to satisfy the *γ*-equation (1), seemingly because, as written above, cases reported now as recovered stopped being contagious some days ago.

The purpose of the current report is to propose techniques to read and interpret the pertinent data, from the point of view of the SIR model represented accurately by equation (1) and roughly by the equation driving new affected cases. The application in mind is to assist in the dynamic assessment of the effective reproduction rate *R*_0_. Some attempts are made to take into account warnings and guidelines, such as those of Holmdahl and Buckee [6]. Performance analysis, validation by simulation and Bootstrap techniques are left as future tasks.

Henceforth, the words “cumulative” and “currently” will be tacitly intended.

## 2 Removed cases - estimation of *γ* and calibration of recovered cases delay

*This Section hinges on the common observation that recovered cases are reported later than dead or affected cases. As a result, the common naive subtraction operation that assesses the current number of infected cases, is positively biased. A method will be presented to gradually shift recovered cases backwards in time to the point that the SIR γ-equation is properly satisfied. At this point the number of infected cases will be minimally perturbed so that the SIR γ-equation is perfectly satisfied. This will provide an assessed day by day number of infected and removed cases, and an estimate of γ. The balance between dead and recovered cases will be ignored along the procedure, but monitored on the end result. The output of this procedure, with the original empirical cumulative number of affected and dead cases and the modified number of recovered cases, is the input for the next Section, dealing with monitoring R*_0_

### Exact solution *Ŷ* to the SIR *γ*-equation

Recalling that *R* = *X − Y*, a perfect match of the SIR *γ*-equation would entail (by adding (1) as a telescopic sum)

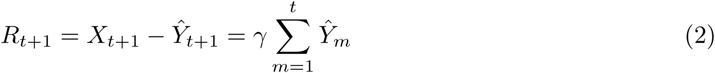

or equivalently

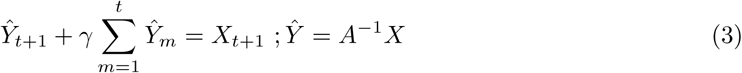

where *A* = *I* + *γB* and *B* is the lower triangular matrix with zeros on and above the diagonal and ones below the diagonal.

The identity in the RHS of (3) is far reaching if justified empirically. It states that if we knew the value of just one number *γ*, the daily number *Ŷ*_*t*_ of currently infected cases could be derived from the cumulative number *X*_*t*_ of affected cases. Furthermore, letting Δ*X*_*t*_ be a smooth version of the daily increase in affected cases (such as a week-long moving average to circumvent missing data on weekends), the effective reproduction rate *R*_0_ could be assessed as

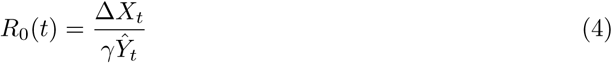

The parameter *γ* can be estimated by minimizing some distance between empirical *Y* and inferred *Ŷ*. This distance will be taken as the *χ*^2^-type distance 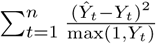. Estimation will be restricted to countries with reliably reported recovered cases. As will be seen, the joint effect of *γ* and *Ŷ* is such that a relatively ample range of *γ* values provide essentially the same assessed process *R*_0_. Hence, the imposition of a central value of *γ* obtained from countries with reliable data (such as Argentina, Brazil, Germany, Israel and Switzerland), will allow the assessment of *R*_0_ in other countries as well (such as Belgium, France, Italy and USA).

### *δ* - late report of recovered cases

A plot of *Y* and *Ŷ* reveals that the two peak at different times. The report of recovered cases seems to be delayed more than the other statistics. In the absence of a model, the parsimonious initial working paradigm is that this additional delay is a constant number of days *δ*. In other words, the effective number of removed cases 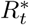 until date *t* is to be taken as the number of dead cases until date *t* plus the number of recovered cases until date *t* + *δ*. The effective number of infected cases 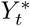 at date *t* is then 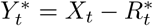. The parameters *γ* and *δ* are jointly determined by minimizing 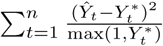.

Once it is realized that shifting in time the number of recovered cases markedly improves the SIR *γ*-equation fit, perhaps it is possible to attempt a gradual shift in time, in case the recording of recovered cases turns progressively more efficient. Such a time delay, in which *δ* is a non-increasing function of time, is described in Appendix 1.

Figure 1 illustrates the results of this method on the Covid19 data from February 22, 2020 to October 18, 2020 in Israel. *γ* was estimated to be 0.0713 allowing the *δ*-delay changes, and 0.0616 on the raw recovered cases. The *R*_0_ process is practically identical in the two estimation modes, and constitutes a smooth alternative to the commonly evaluated empirical *R*_0_-curve 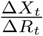 (yellow). The number of infected cases *Ŷ* is practically identical to the delay-modified *Y* but at some distance to the raw *Y*. I.e., the time-delay modification of the recovered cases brings the SIR *γ*-equation (1) to be almost exactly satisfied. Conveniently, the determination of *R*_0_ is insensitive to the time-delay modification. Israel went through a period with *R*_0_ below 1 from day 80 (April 10) to day 128 (May 28) and then steadily above 1 until day 255 (October 2), with a period with *R*_0_ = 1 between day 195 (August 3) to day 212 (August 20). *R*_0_ has been below 1 and decreasing since day 255 (October 2). The empirical yellow *R*_0_ curve tells the same story, more noisily. The empirical analysis assessed higher numbers of infected cases in the first wave and lower numbers towards the end of the monitoring period. The empirical data identified a middle wave with more infected cases, where the time delay method kept *R*_0_ steady at 1.

**Figure 1:**
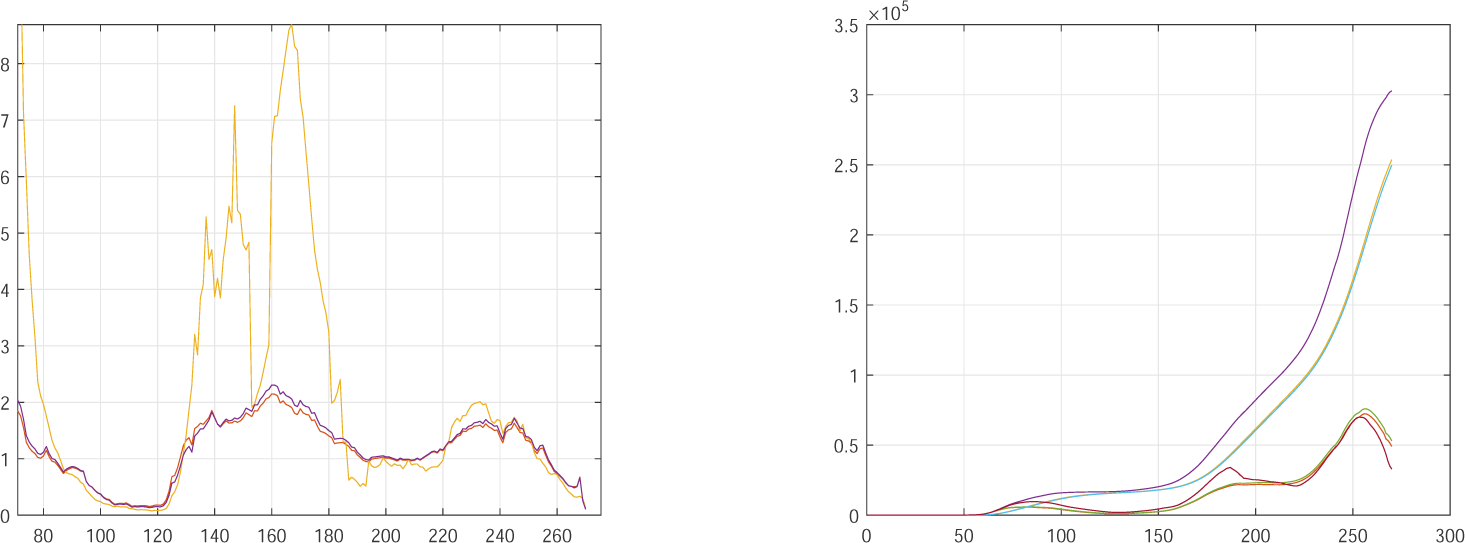
Israel Covid19, February 22 to October 18, 2020. Left: Empirical *R*_0_ (yellow) and derived *R*_0_, with (*γ* = 0.0713) and without (*γ* = 0.0616) delay. Right: affected, removed and infected cases - the latter empirical in red, with and without delay.

Whichever method is used, raw or time delay, the estimates of *γ* (see Table 1) differ too much from country to country. The reaction to such diversity is presented as the following exercise, motivated by the inescapable observation that some countries under-report the number of recovered cases.

**Table 1:** Estimation of the parameter *γ*, raw data and time delay. January 22 to October 18, 2020 (Brazil until October 9).

| Country | Argentina | Belgium | Brazil | France | Germany | Israel | Italy | Suisse | USA |
| --- | --- | --- | --- | --- | --- | --- | --- | --- | --- |
| raw $\gamma$ | 0.0704 | 0.0051 | 0.0705 | 0.0062 | 0.0687 | 0.0616 | 0.0325 | 0.0695 | 0.0083 |
| delay $\gamma$ | 0.0711 | 0.0052 | 0.0713 | 0.0064 | 0.0759 | 0.0713 | 0.0404 | 0.1233 | 0.0084 |

Exercise: The parameter *γ* is fixed as 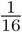. The data on recovered cases is ignored, and the number of infected cases *Ŷ* is derived from the number of affected cases only, by means of formula (3).

Figure 2 Left illustrates the evaluation of *R*_0_ in USA. Empirical (yellow) and derived curves (purple) as if data was complete, and derived curve for 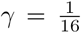 (red). Figure 2 Center displays affected, recovered and infected cases as assessed under 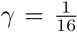. According to this analysis, USA had *R*_0_ below 1 from day 105 (May 5) to day 145 (June 14) and from day 193 (August 1) to day 240 (September 17). *R*_0_ has been above 1 and steadily increasing since day 247 (September 24). Crossovers from above to below 1 can be identified directly from *R*_0_ or as local maxima of the number of infected cases (red curve in Figure 2 Center). Figure 2 Right displays nonsensical straightforward raw data.

**Figure 2:**
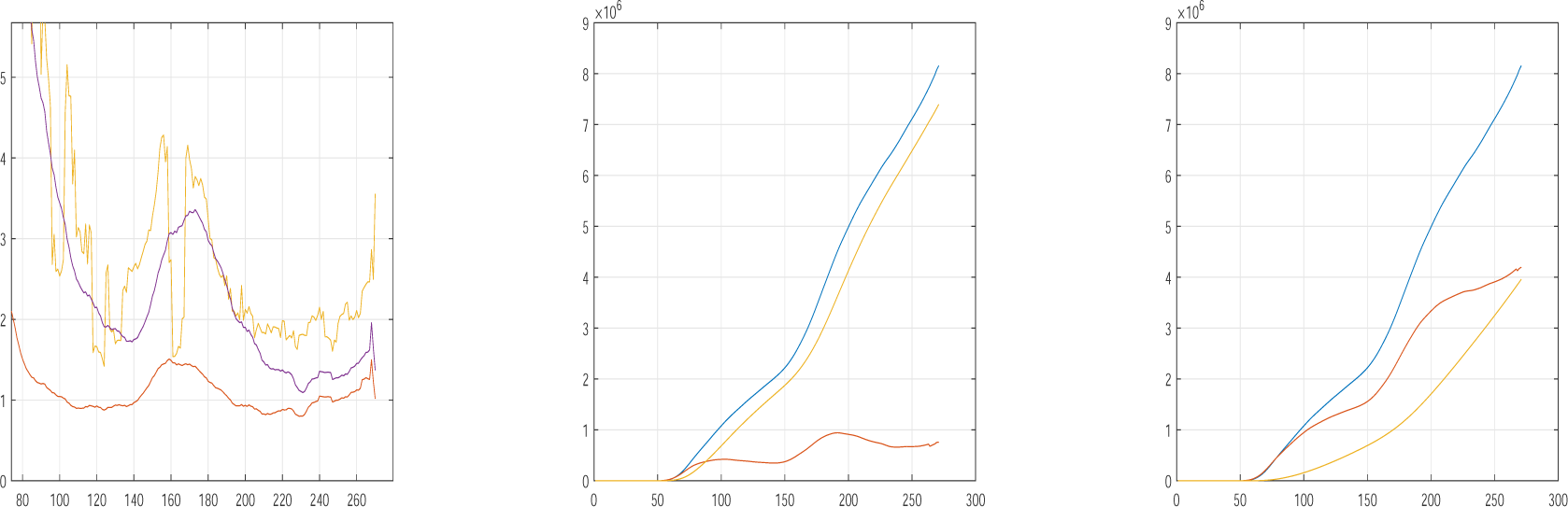
USA Covid19, February 22 to October 18, 2020. Left: Empirical *R*_0_ (yellow) and derived *R*_0_ for estimated *γ* = 0.0084 (purple) and for *γ* set as 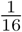 (red). Center: affected (blue), removed (yellow) and infected (red) cases under *γ* set as 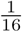. Right: affected (blue), removed (yellow) and infected (red) cases under estimated *γ* = 0.0084.

Figure 3 displays for Belgium (left), France (center) and Italy (right) the findings parallel to those displayed for USA in Figure 2Left. The (most optimistic but deemed the only realistic) red curves place *R*_0_ steadily above 1 from day 220 (August 28) in Belgium, from day 160 (June 29) in France and from day 180 (July 19) in Italy. The value is assessed on October 18 as 1.5 in France and well above 2 in Belgium and France.

**Figure 3:**
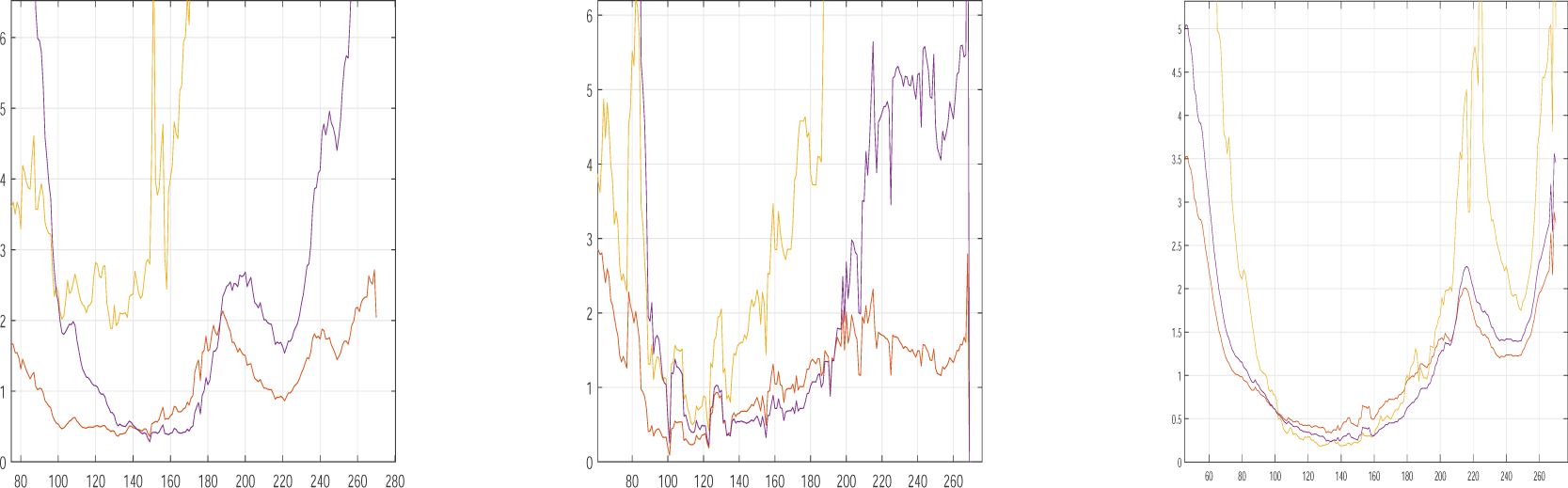
Covid19, February 22 to October 18, 2020. Empirical *R*_0_ (yellow), raw derived *R*_0_ (purple) and 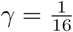-derived *R*_0_ (red) are different in Belgium (left), France (center) and Italy (right).

What follows is a comparison with countries that seem to measure recovered cases more accurately. Figure 4 displays under estimated *γ* for Argentina Brazil, Germany and Switzerland the findings parallel to those displayed under *γ* fixed at 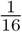 for USA in Figure 2 Left and Belgium, France and Italy in Figure 3. The empirical blue curves are a noisy version of the derived red curves. Argentina and Brazil present a similar behavior, with Brazil recovering faster from the epidemic. Germany and Switzerland present similar behavior to the European countries displayed in Figure 3. This lends credibility to the assessment of *R*_0_ by setting a central value of *γ* when recovered cases ought to be ignored.

**Figure 4:**
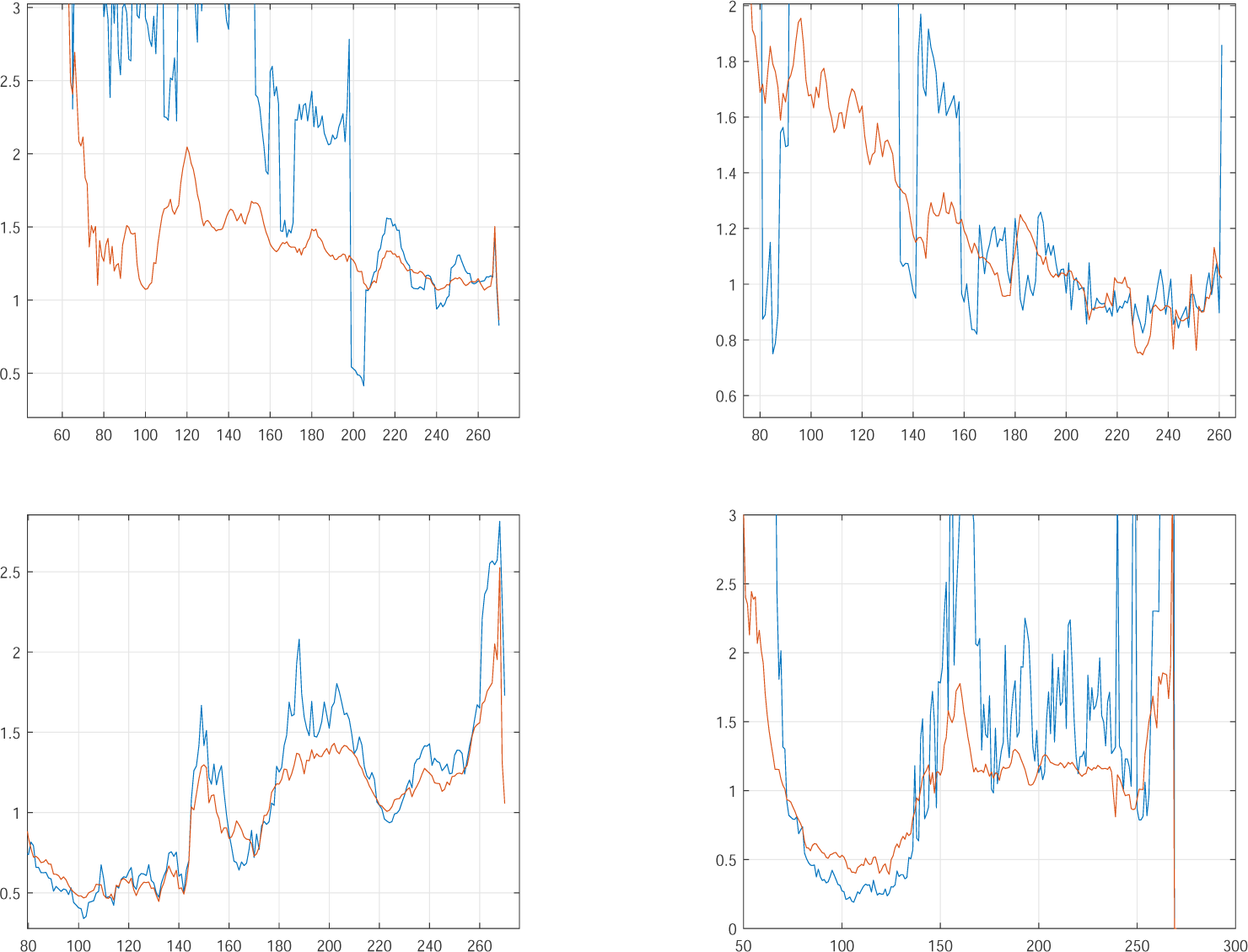
Covid19, February 22 to October 18, 2020. *R*_0_ derived under estimated *γ* (red) versus empirical *R*_0_ (blue) are similar in Argentina (top left), Brazil (top right), Germany (bottom left) and Switzerland (bottom right). The derived version is more stable.

## 3 Instability of *γ*: the deficient report of recovered cases

Figure 5 Left displays the horizontal distance between the affected and removed curves in Israel as presented in Figure 1, analyzed with and without time delay. It is apparent that the two curves are roughly parallel, at distance roughly 20 days, consistent with the estimation of *γ* as being between 0.06 and 0.065. Figure 5 Right displays in blue color the horizontal distance between the affected and removed curves in the USA data, for the estimate 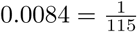 of *γ*. USA heavily under-reports the number of recovered cases, and these impossible findings are the result of taking the data on face value. Figure 5 Right displays in red color the horizontal distance between the affected and recovered curves in the USA data for *γ* set as 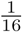 in the assessment (3) of the infected cases *Ŷ*. This interpretation of infected cases allows the possibility of estimating the *R*_0_ curve for countries that under-report recovered cases. USA is displayed in Figure 2 Left, Belgium, France and Italy are displayed in 3.

**Figure 5:**
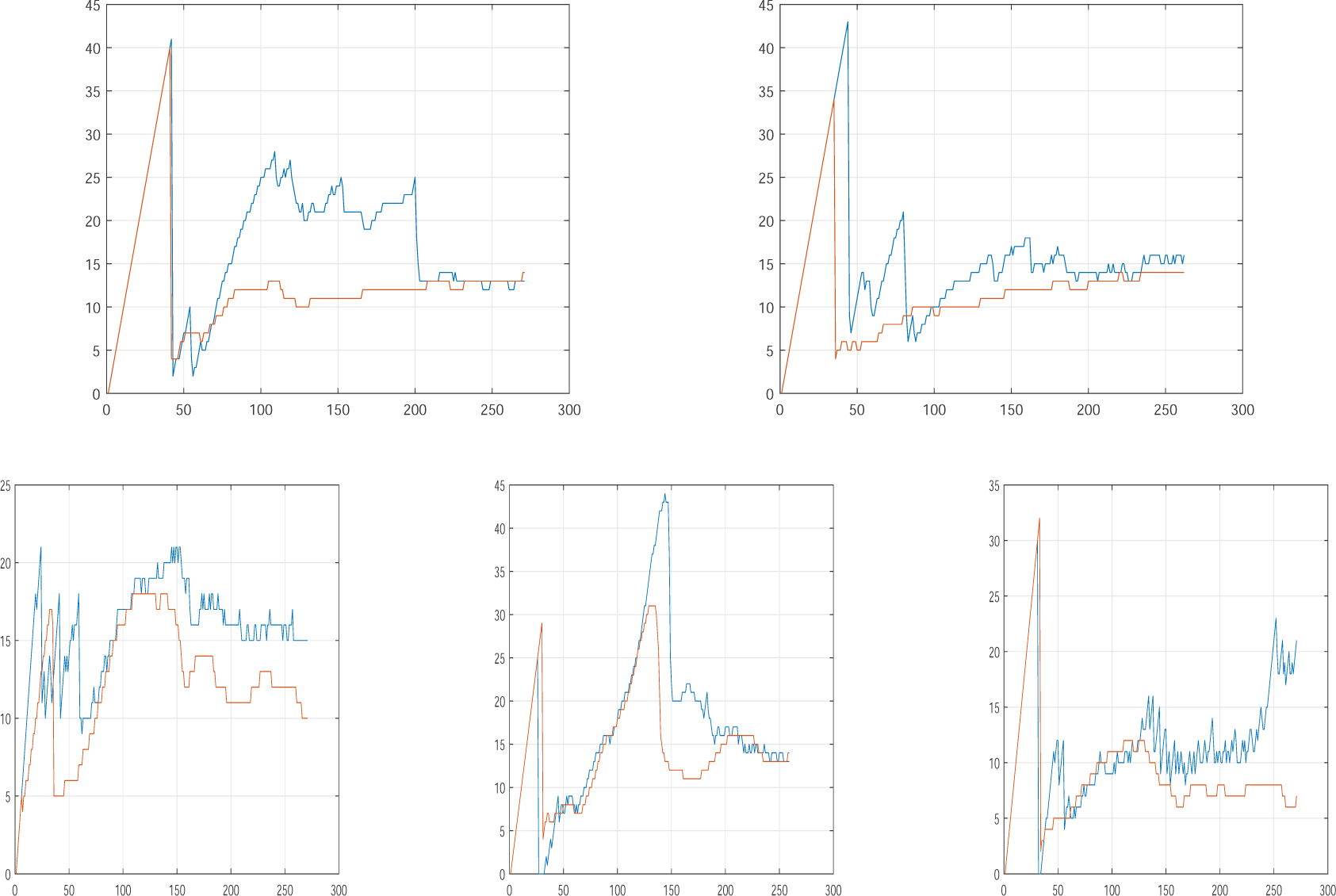
Horizontal distance between the affected and removed curves, January 22 to October 18, 2020. Raw (blue) and derived (red) are similar. Top left: Argentina. Top right: Brazil. Bottom left: Germany. Bottom center: Israel. Bottom right: Switzerland.

As mentioned in the Introduction, the meaning of *γ* is perhaps carried more crisply by 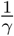, the theoretical average time an infected individual is contagious until removed (recovered or dead). Table 1 reports values too different for *γ* in the various countries illustrated. It is inconceivable that this average could be over 100 days in Belgium, France and USA, 30 in Italy on the one hand, and around 15 days in the other countries. The horizontal distance between the affected and removed curves in Figures 1 and 2 should in principle be this average infective time 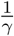, the time it takes from the moment the number of affected cases reaches some height until an equal number of cases has been removed. This seems correct in Israel but not in USA.

Figures 5 and 6 displays these horizontal distances. In a nutshell, the horizontal distance should be roughly 16 days. In periods between waves of the epidemic, when the number of affected cases stays roughly constant, this distance may be ill defined, with a tendency to appear bigger. But all in all, it should be roughly constant, as in Argentina, Brazil, Germany, Israel and Switzerland. However, a sharply increasing horizontal distance indicates under-reported recovered cases. This is apparent in Belgium, France, Italy and USA.

**Figure 6:**
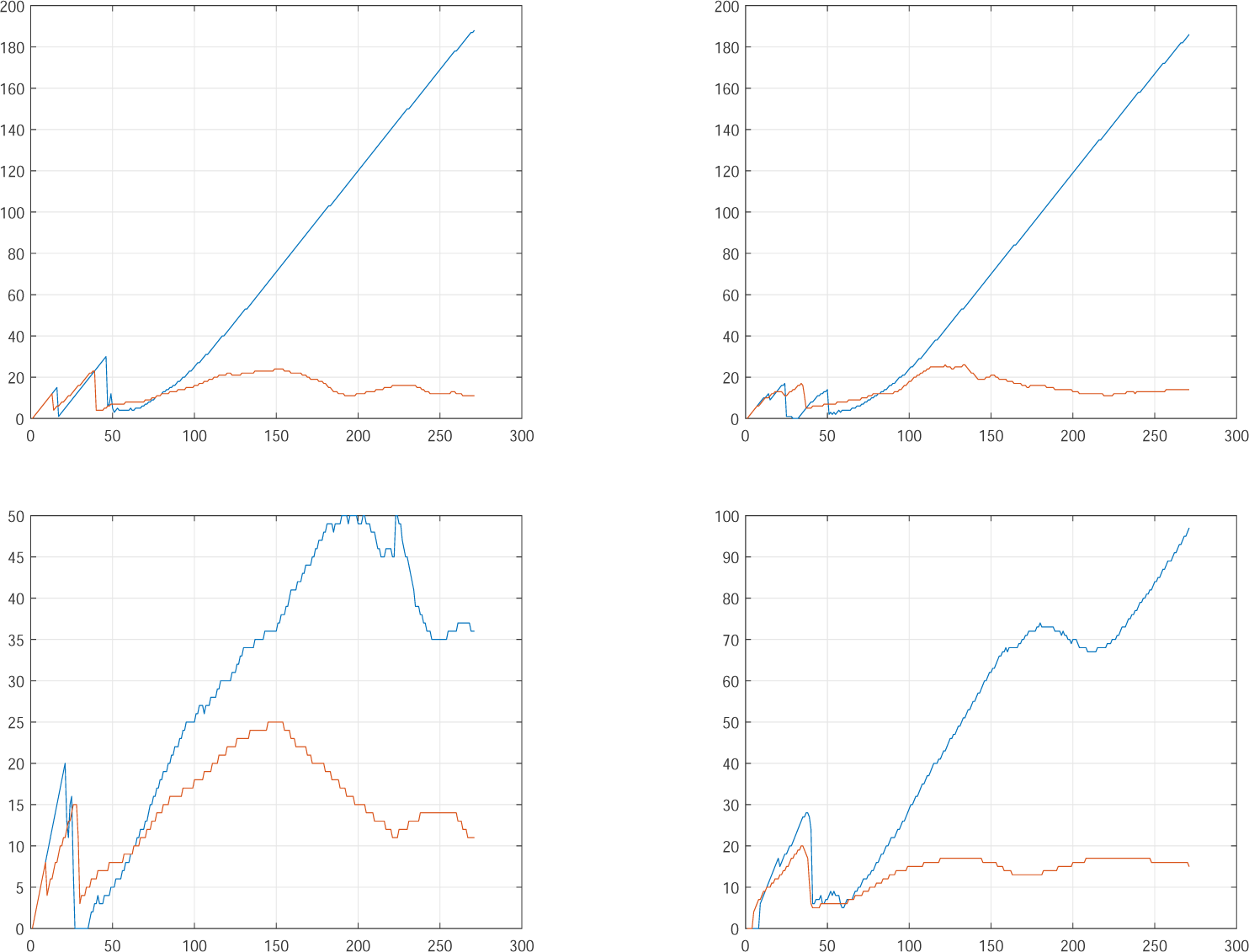
Horizontal distance between the affected and removed curves, January 22 to October 18, 2020. Raw (blue) and derived (red) are different. Top left: Belgium. Top right: France. Bottom left: Italy. Bottom right: USA.

## 4 Removed cases divided into recovered and dead cases

This division is not part of the standard SIR equations. However, it is important on its own sake, and can shed some light on data quality. Figure 7 displays the proportion of incremental dead cases out of incremental removed cases, smoothed by a two-week-window moving average. As a rule, all countries displayed show a sharp decrease of this proportion along time. USA shows a decrease from 40% to 3.5% with *γ* estimated as 0.01. The parallel decrease in Germany and Switzerland is from 4% or 5% to less than 1%, with respective *γ* estimated as 0.085 and 0.125. Israel shows a decrease from 2.5% to less than 1%, with *γ* estimated as 0.056. Brazil and Italy stand in between, with *γ* assessed respectively as 0.064 and 0.037 and lethality proportion decreasing from 10%-20% to 2%.

**Figure 7:**
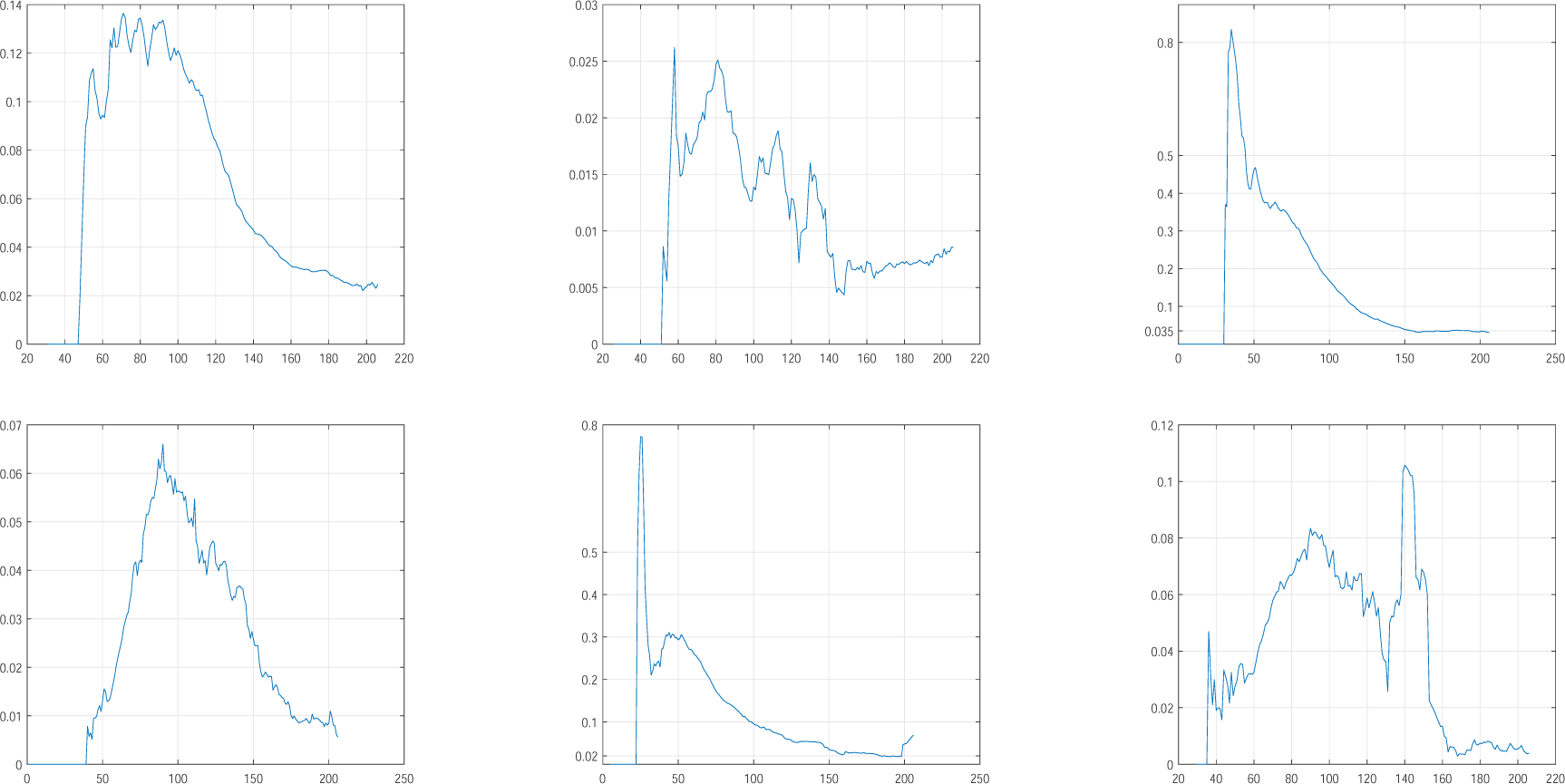
The proportion of incremental dead cases out of incremental removed cases. From top left to bottom right: Brazil, Israel, USA, Germany, Italy, Switzerland.

This disparity may suggest that the identification of Covid19 as infection and cause of death differ markedly between countries. This issue needs attention.

## 5 Conclusions

Analysis of Covid19 dynamics has been performed on and restricted to the most reliable publicly available data, the data repository at Johns Hopkins University. Evidence has been found that recovered cases are only partially recorded in some countries, and the delay in recognizing recovered cases seems longer than the recognition of affection or death. Attempts to adjust the data so as to correct effects of this delay, have led to date-aligned data that satisfies the relation between removed and infected cases claimed by the SIR *γ*-equation in epidemiology (Kermack and McKendrick [7]). This correction method has been applied to assess missing recovery data and allow monitoring of the effective reproduction rate *R*_0_.

The methods presented have been illustrated on the data of each of nine countries, five of which with seemingly reliable recovery data.

## Data Availability

The data analyzed in this work is taken from the COVID-19 Data Repository by the Center for Systems Science and Engineering (CSSE) at Johns Hopkins
University.

## 6 Appendix 1: Variable time delay of recovered cases

The number of recovered cases *REC* displays occasionally sharp jumps. Suppose that on a particular day the number of newly recovered cases is five times the daily average in the next week. It makes sense to shift the number of recovered cases (by four or less days) backwards in time before this date rather than globally. This procedure is performed in small steps, as follows. Let Δ*REC*_1_(*t*) be the number of new recovered cases in day *t* and let Δ*REC*_2_(*t*) be the daily average of new recovered cases in the week following day *t*. Suppose that the ratio 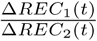, taken from day 40 or 50 to a week before the end of observation, is maximal at day *t*_1_, where it exceeds some threshold like *TH* = 3. The function *RC*(*t*) will be modified as follows: It stays as is from day *t*_1_ onwards, it is shifted backwards by one day before day *t*_1_, and the single day left unassigned is given as value the simple average of the two days next to it. The procedure is applied again and again to the newly defined function, until no ratio exceeds the threshold, or some maximal number of steps, such as 5 or 10, has been performed. The rationale behind choice of *TH* is that after adding a day when the ratio is *TH*, the new ratio will be 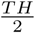. Hence, *TH* should be in principle somewhere between 2.5 and 3. Since data has been smoothed by week-long moving averages, the choice of *TH* could be smaller. The choice *TH* = 2.5 is adopted.

If the maximal allowed number of steps is kept small, the modified vector of numbers of recovered cases can be then submitted to the global *δ* procedure to account for a possible delay towards the end of the observation period.

## 7 Appendix 2: *R*_0_ in some of the states of the USA

Due to under-reporting of recovered cases, no attempt can be made to estimate *γ*. The central value 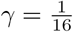 is adopted, and the number of infected cases is derived from (3). The effective reproduction rate *R*_0_ is then calculated via (4). These assessments are displayed in Figure 8 for a choice of twelve states of the USA. *R*_0_ has an increasing trend towards October 18 in all of these states, for all of which *R*_0_ either exceeds or approaches 1 at this date.

**Figure 8:**
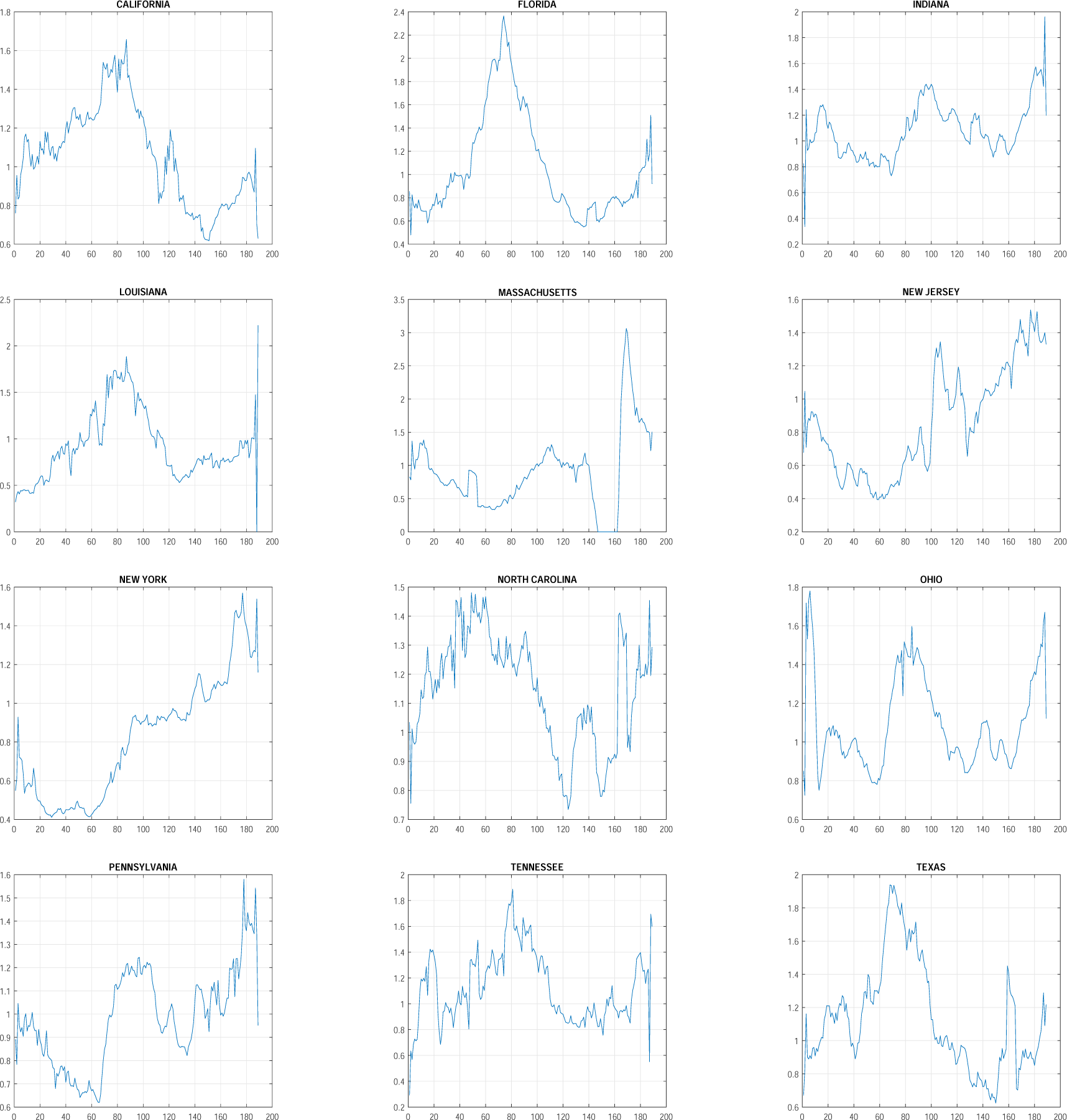
Covid19, April 12 to October 18, 2020. *R*_0_ derived under 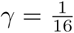 for a choice of states of the USA.

## Acknowledgements

Thanks are due to Nitay Alon, Yoav Benjamini, Peter Bickel, Ilan Eshel, Eytan Ruppin, Laura Sacerdote, Dov Schvarts and David Steinberg for helpful suggestions.

The data analyzed in this work is taken from the COVID-19 Data Repository by the Center for Systems Science and Engineering (CSSE) at Johns Hopkins University.

